# Reproducibility of electroencephalography biomarkers for diagnosis of major depressive disorder

**DOI:** 10.1101/2024.11.11.24317109

**Authors:** Yasmin Hollenbenders, Friedrich Ph. Carrle, Roman Mähler, Christoph Maier, Alexandra Reichenbach

## Abstract

Major depressive disorder (MDD) and other psychiatric diseases can greatly benefit from objective decision support in diagnosis and therapy. Machine learning approaches based on biomarkers extracted from electroencephalography (EEG) have the potential to serve as low-cost decision support systems. Although this approach has shown promise, inconsistent findings regarding the diagnostic value of those biomarkers impede their clinical translation. Therefore, the *replicability* and *robustness* of these biomarkers need to be established first. We employ a multiverse analysis to systematically investigate the *robustness* against variations in six data processing steps, which may be one source of contradictory findings. These steps are artifact removal, normalization, time-series segment length, biomarker from the alpha band, aggregation, and classification algorithm. To ensure *replicability* of our findings, we analyze two publicly available EEG datasets with eyes-closed resting-state data containing 25/18 MDD patients and 23/14 healthy control subjects. The accuracies of diagnostic classifiers range from 81% to chance level, dependent on dataset and combination of processing steps. We observe that the selection and combination of processing steps significantly influences the results. Overall, the *replicability* of our findings across the two datasets was inconsistent. This study is a showcase for the advantages of employing a multiverse approach in EEG data analysis and advocates for larger, well-curated datasets to further neuroscience research that can be translated to clinical practice.

## Introduction

Major depressive disorder (MDD) is the most common mental disorder with a worldwide lifetime prevalence of 10.6%.^1^ However, epidemiologic data suggests that a substantial number of cases is unreported, which might be due to current diagnostic practices, stigmatization, or public health policies.^2^ In order to advance knowledge of the disorder, identify its biological underpinnings, improve diagnostic procedures, and develop more effective treatments, biomarkers are needed. Electroencephalography (EEG) constitutes a non-invasive and cost-efficient method to provide such biomarkers by recording brain physiology. Indeed, studies have shown that biomarkers extracted from EEG signals can distinguish between MDD patients and healthy controls (HC).^3–5^ However, results contradict each other regarding discriminatory features extracted from these signals.^3,5^ The heterogeneity of the disorder as well as methodological differences across studies likely contribute substantially to these inconsistent findings,^5,6^ and complicate reproducibility.

Resolving these contradictions has increasingly moved into the focus of neuroscience research by promoting the importance of reproducibility. Botvinik-Nezer et al.^7^ distinguish three complementary dimensions of reproducibility, two of which are applied in this study. *Replicability* captures whether implementing the same analytical approach to different datasets yields comparable results. *Robustness* to analytical variability characterizes the stability of findings for the same data across different analytical approaches. Variability can, therefore, enter the data-processing pipeline through decisions made at multiple stages. Choices about population selection, study design, and data acquisition, including technical factors, can affect *replicability*, whereas decisions about preprocessing, feature extraction, and the final distinction between MDD and HC may challenge the *robustness* of findings.

*Replicability* is influenced by characteristics of data collections, including but not limited to participant selection, diagnostic assessment and labeling, symptom documentation, sample size, and electrode placement. These parameters are fixed once data have been recorded and, therefore, cannot be modified when working with publicly available open datasets. Nevertheless, *replicating* findings using public data or newly collected samples, in combination with the original processing pipeline, remains essential for corroborating findings.^7^

Once data is recorded, preprocessing steps such as filtering, artifact removal, normalization, segmenting, subsampling or augmentation, and data aggregation are often applied to the EEG signal. Although this sequence represents commonly used steps, the selection of steps and the specific methodological choices within each step can vary considerably, potentially leading to divergent findings and challenging *robustness*.^8^ Artifact removal is a central step in preprocessing EEG data but the appropriate degree of intervention during this process is discussed highly controversial. Many studies rely on fully automated^9^ or (semi-)manual cleaning procedures,^10,11^ whereas other researchers argue for minimizing artifact removal to preserve more of the original neural activity.^12^ Along the same line, normalization approaches range from omitting normalization entirely,^9^ to applying subject-wise normalization to harmonize datasets,^13^ or to implementing channel-wise normalization in order to stabilize input distributions for machine-learning (ML) models.^14^ Each strategy embodies distinct assumptions about inter-subject variability and model sensitivity. Furthermore, segmentation of EEG time-series is frequently employed in ML-based EEG analysis to increase the amount of training data. Here, the segment length can significantly influence model performance^15^, highlighting the importance of carefully selecting temporal windows. Finally, while ML approaches generally benefit from larger datasets, statistical analyses often take the opposite route by aggregating data to robust estimators to reduce noise and emphasize stable patterns.

Alpha waves are associated with relaxation and predominantly observed when eyes are closed.^16^ Biomarker from the alpha band are as prominent in MDD research^3^ as they are controversially discussed.^17^ While some studies suggest that biomarkers from the alpha band perform better than markers from other frequency bands,^18,19^ other studies found that they perform worse.^15,20,21^ Furthermore, it is disputed whether alpha activity is higher or lower in MDD subjects compared to HCs.^18,19^ Yet, all these studies used different processing methods, rendering their results non-comparable.^17,22^ Even though the alpha band is one of the most consistently defined frequency bands, usually specified from 8 to 13Hz, its definition can range anywhere within the borders of 6 to 14 Hz, introducing one source of analytical variability.^6^ Additionally, biomarkers sharing the same name are calculated in different ways such as the frequently reported alpha asymmetry,^23^ further hampering reproducibility.^7^ To ensure the reliability of findings regarding the neurophysiological underpinnings of MDD and their deviation to the neurophysiology of HC, objective biomarkers not only with high discriminatory power but also *replicability* across datasets, and to some degree of *robustness* to analytical variability are required.

The analytical approach for differentiating between patients and HCs represent another key variability across studies. These differences can be analyzed, e.g., with statistical comparisons, or with ML algorithms that constitute an increasingly popular multivariate approach. The latter are frequently applied to mimic a clinical decision-making process by modeling a classification problem with the class labels MDD and HC. The approaches range from classical supervised algorithms that usually operate on hand-crafted biomarkers^24^ to more complex deep learning models that often, but not exclusively, operate on the raw time-series data or their frequency domain representation to inherently derive biomarkers in the training process^4^.

In most studies, the data recording and processing pipeline consists of one specific choice out of several possible alternatives. Yet, different methodical decisions during this process can have an impact on the findings, rendering comparison between studies hard.^24^ One approach to reconcile the different findings and demonstrate *robustness* of the analysis of EEG data to the analytical variability is the multiverse analysis.^25^ This approach has been adopted in applied neuroscience to reconcile contradictory findings about alpha asymmetry in MDD,^26^ or to recommend an optimized processing pipeline.^27^ This analysis strategy enables the demonstration that the high number of degrees of freedom in conducting EEG studies is one cause for the lack of reproducibility in findings.^5,6^

The goal of this study is to assess two types of reproducibility on EEG alpha band biomarkers for the support of MDD diagnosis. In order to mimic a diagnostic scenario yet still retain close connection to studies using statistical analyses on those biomarkers, we adopt a classification approach with classical ML algorithms based on hand-crafted EEG markers. To keep the focus on the analytical variability, we limit our investigation to linear alpha markers, the most frequently discussed category of EEG biomarkers for MDD^3^. For the purpose of *replication*, we utilize the two most frequently used publicly available MDD datasets^28^ with eyes-closed resting-state EEG and harmonize them.^13^ To assess the *robustness* of the biomarkers to analytical variability, we systematically investigate the influence of several preprocessing steps, different features, and classification algorithms by means of a multiverse analysis. This study does not examine assumptions or predictions regarding specific biomarkers.

## Methods

### Data

Two publicly available datasets (D1^29^; D2^30^) with MDD patients (D1: n=29 (presumably age: 40.3±12.9; female: n=17)^31^, D2: n=24 (age: 30.9±10.4; female: n=11)) and HCs (D1: n=28 (presumably age: 38.3±15.6; female: n=9)^31^, D2: n=29 (age: 31.5±9.2; female: n=9)) were downloaded in August and October 2021, respectively. All information about D1 were derived from the original study describing the dataset^31^, because no accompanying information was provided with the dataset. The study designs were approved by the respective ethics committees (D1: Human ethics committee of the Hospital Universiti Sains Malaysia (HUSM); D2: Ethics Committee for Biomedical Research at the Lanzhou University Second Hospital) and participants provided written informed consent. All patients were diagnosed based on DSM-IV criteria. Patients from D2 did not take any anti-depression medication during the two weeks before recordings were taken. Both datasets contain 5-minute resting-state EEG time-series data with eyes closed. D1 was recorded with 19 electrodes placed according to the 10-20 system^32^ with linked-ear-reference and 256 Hz sample frequency. D2 was recorded with 128 electrodes in the Geodesic Sensor Net^33^ with Cz as reference electrode and 250 Hz sample frequency.

### Dataset Harmonization

To establish a common ground for the analyses and limit preprocessing variability, we harmonized the datasets as far as possible using the Python Package *MNE*^34^ (version: 1.2.3). We matched the 128 electrodes from D2 to correspond to the 10-20 system according to the supplementary material provided with the dataset. Only data from the 13 overlapping electrode locations was included in the analysis: seven (pre)frontal (Fp1/2, F3/4, F7/8, Fz), two central (C3/4), two parietal (P3/4), and two occipital (O1/2). Afterward, both datasets were re-referenced to common average, a prerequisite for the independent component analysis (ICA) in the cleaning step. D1 was resampled to 250 Hz to match the sampling rate of D2. To harmonize the spectral bandwidth, both datasets were bandpass filtered from 1 to 40 Hz to include the filters originally applied to the two datasets.

### Multiverse Analysis

We adapted a multiverse analysis^25^ to investigate the impact of processing choices on the classification results. For each step (for analysis purposes also referred to as factor) of the processing pipeline, we implemented different options (also referred to as factor levels) and combined them fully. This resulted in a multiverse comprising 4752 parallel processing paths (Table 1) for each dataset. Note that the chosen processing steps and variations are only a selection of possible options and not an extensive list.

**Table 1:** Processing steps / factors and their variations / factor levels included in the multiverse analysis.

| **Processing Stage** | **Processing Step / Factor** | **Variations / Factor Levels** | **Multiverse Analysis Conditions**<br>**[# paths total]** |
| --- | --- | --- | --- |
| Cleaning | artifact removal | none; automatic; manual revision | 3 [3] |
| Preprocessing | normalization | none; subject-wise; channel-wise | 3 [9] |
|  | butterworth filter | 4<sup>th</sup> order; 8<sup>th</sup> order | 2 [18] |
|  | segment length | 5s; 10s; 15s; 20s | 4 [72] |
|  | remove segments with artifacts (flagged by manual revision) and subsampling of the segments (for all paths) |  |  |
| Feature Extraction | biomarker | 11 alpha band (8-13 Hz)<br>biomarker (cf. Table 2) | 11 [792] |
|  | aggregation | none; median over 8 segments | 2 [1584] |
| Classification | algorithm | Support Vector Machine (SVM);<br>Logistic Regression; Random Forest | 3 [4752] |
|  |  |  | **4752 paths** |

### Cleaning

EEG data is susceptible to environmental, technical, and physiological interference.^35^ Three approaches for artifact removal were carried out: 1) No artifact removal^12^. 2) Fully automatic removal of artifact components: EEG signals were separated into components with an ICA^36^, automatically labeled with *ICLabel*^37^ (version: 0.4), and reconstructed without the components affected by artifacts, i.e., eye blinks, muscle artifacts, heartbeat, line noise, and channel noise. 3) Customized artifact removal with manual revision: An ICA was performed to identify common artifacts like eye and muscle movement as well as low-frequency anomalies and dataset specific technical artifacts. Identified artifact components were removed or attenuated using wavelet denoising. The pre-cleaned data was manually revised to exclude remaining high and low frequency artifacts by flagging affected time frames.

### Preprocessing

Normalization is applied to EEG signals in order to level amplitudes and make subjects more comparable.^4^ To compare the impact of normalization methods, we applied three variations of *z*-normalization: 1) No normalization. 2) Subject-wise normalization is applied to level putative differences across subjects. For this, all channels of each subject are *z*-normalized jointly to align only the subjects of the datasets with each other while retaining the between-channel differences for each subject. 3) Channel-wise normalization is frequently used to level putative differences across EEG traces. Each channel of each subject is *z*-normalized separately. Note that channel-wise normalization destroys spatial information based on absolute values of the power of the signal. Furthermore, biomarkers based on distribution, a certain frequency, or calculated relating to the full band are not affected by any of the normalization methods.

After normalizing, we split the data into non-overlapping segments of 20s. All segments containing flagged artifacts were removed. This procedure was applied both to the cleaning path with manually revised data as well as to the paths without and with automatic artifact removal. This balances the comparability across datasets with a putative bias introduced by this approach. We selected the number of segments to maximize both the amount of usable data and the number of remaining subjects, resulting in eight randomly subsampled segments per subject. Subjects with less than eight remaining segments were excluded from subsequent analyses. This left 48 (MDD: 52%) / 32 (MDD: 56%) subjects for D1/D2. Splitting recordings into segments of the same duration is a common method to augment data, yet there is no best practice on how long these segments should be and reported time spans vary widely. Therefore, we additionally considered the first 5, 10, and 15 seconds of each segment for the analysis.

### Feature Extraction

To compile a comprehensive set of alpha band biomarkers, we calculated eleven spectral biomarkers common for diagnosis of psychiatric disorders (Table 2).^3^ We limited the analysis to biomarkers that can be calculated individually for each electrode resp. EEG channel to avoid combinatoric explosion of the number of possible features. The frequency range was set to 8 to 13 Hz, the typical range of the alpha band.^6^ To obtain the spectral power characteristics of the signal, we used Welch’s method with Hanning window, 50% overlap, and a window length of 512 data points. This results in 2.05s window duration and a spectral resolution of 0.5 Hz. To calculate the envelope of the signal, we first extracted the alpha band with a Butterworth bandpass filter of either 4^th^ or 8^th^ order. The envelope biomarkers were calculated based on the envelope of the time-domain signal,^38^ which was calculated using the absolute values of the Hilbert transform. The actual biomarkers characterize the distributions of the envelope data points. Because the calculation of some of the biomarkers is inconsistent across studies or not properly described in the literature, we compiled a detailed overview of their construction to enable analytical reproducibility (Table 2). Biomarkers were calculated using the Python toolboxes *scipy*^39^ (version: 1.9.3), *numpy*^40^ (version: 1.23.5), and the function *bandpower* adopted from Vallat and Walker^41^. The accompanying source code for biomarker calculation further ensures analytical reproducibility (see supplementary files “calculate_biomarkers.ipynb” and “alpha_markers.py”).

**Table 2:** Biomarkers used in this study, their description, calculation, and sources/studies where those biomarkers were used for diagnosis or characterization. * indicates biomarkers that are invariant to normalization.

| **Biomarker** | **Description** | **Used In** |
| --- | --- | --- |
| absolute band power | Total power of the alpha band.<br>Welch periodogram, and integration using Simpson’s rule (adopted from <sup>41</sup>) | 15,18–21 |
| relative band power* | The absolute power of the alpha band divided by the absolute power of the EEG signal (i.e., from 1 to 40 Hz) (adopted from <sup>41</sup>) | 20 |
| absolute centroid frequency* | Center of mass of the alpha band power spectrum.<br>Welch periodogram, and calculation of the power-weighted sum of the frequencies of the alpha band | 15,20 |
| relative centroid frequency* | Absolute centroid frequency of the alpha band divided by the absolute centroid frequency of the total EEG band (here: from 1 to 40 Hz) | 20 |
| peak frequency* | The frequency of the alpha band spectral component with the highest power.<br>Welch periodogram, select frequency with maximum power<sup>42</sup> | 15,20,38 |
| envelope kurtosis* | Butterworth bandpass filter, Hilbert transform, and calculating the kurtosis of the absolute values of the transformed signal | 38;<br>on raw signal:<br>20 |
| envelope skewness* | Butterworth bandpass filter, Hilbert transform, and calculating the skewness of the absolute values of the transformed signal | 38;<br>on raw signal:<br>20 |
| envelope median | Butterworth bandpass filter, Hilbert transform, and calculating the median of the absolute values of the transformed signal | 38 |
| envelope interquartile range | Butterworth bandpass filter, Hilbert transform, and calculating the interquartile range between the 25<sup>th</sup> and the 75<sup>th</sup> quartile of the absolute values of the transformed signal | 38 |
| envelope variance | Butterworth bandpass filter, Hilbert transform, and calculating the variance of the absolute values of the transformed signal | <sup>38</sup>;<br>on raw<br>signal:<br>20 |
| envelope range | Butterworth bandpass filter, Hilbert transform, and calculating the range of the absolute values of the transformed signal | <sup>38</sup> |

Biomarker values were calculated individually for each time segment and EEG channel. Since there are eight segments per subject, one subject is represented with eight different values, which effectively corresponds to a data augmentation process commonly encountered in the field of ML. Alternatively, we pursued an aggregation strategy by taking the median over each subject’s values, resulting in one single and putatively more robust biomarker value per subject and EEG channel. The latter is a typical approach for analyses with statistical inference tests.

### Classification

The two datasets were used to train individual classification models with three commonly used and simple classification algorithms well suited for smaller datasets: Logistic Regression (maximum 200 iterations), Support Vector Machine (SVM) with a linear kernel (default values), and Random Forest (maximum depth 2). Models were trained for each individual biomarker. For each model, the input was a 13-dimensional feature vector containing the biomarker value for each EEG channel. The class labels were the diagnostic groups MDD and HC. The models were trained with six-fold cross-validation stratified by diagnosis on a per-subject split basis^13^, resulting in five to eight subjects per test dataset. To counteract imbalance of the class distribution, the balanced mode was used. Note that the partitioning in training and test dataset was kept identical for all paths of the multiverse analysis to ascertain comparability. Balanced accuracy of each model was used for the analysis. We used *sklearn*^43^ (version: 1.1.3) for all classification-related implementations.

### Analytical Approach

In this study, we assess two types of reproducibility^7^, both from different perspectives. The *replicability* was assessed between as well as within datasets. *Replication* between datasets is assessed by using two publicly available datasets and observing convergences and divergences between those across all further *robustness* analyses. *Replication* within datasets is analyzed by using six-fold cross validation and comparing results between folds. *Robustness* is assessed for both datasets regarding processing steps and biomarkers. To assess the *robustness* of the classification performance against variations in the processing steps, we conducted analyses of variance (ANOVAs) including the factors of the multiverse as within-subject factors and subsequent ANOVAs or *t*-tests for post hoc analyses whenever appropriate. We categorized the ANOVAs into two groups of biomarkers: those affected by normalization (A-BM: **envelope median**, **range**, **variance**, **interquartile range** and the **absolute band power**) and those unaffected by normalization (U-BM: **absolute** and **relative centroid frequency**, **envelope skewness** and **kurtosis**, **relative band power**, and **peak frequency**). ANOVAs for U-BMs are conducted without the normalization factor. The *robustness* of the individual biomarkers was further assessed with three approaches: a) Analysis of the performance range of each path for each biomarker. b) One-tailed *t*-tests against chance level (i.e., 50% accuracy) to test each path for above chance level classification performance. c) Correlation analysis between paths with biomarker as variables to analyze biomarker patterns across paths.

Additional analyses include Fischer’s exact test to compare groups with nominal scaled data. ANOVAs were conducted with *statsmodel*^44^ *AnovaRM* (version: 0.14.0), post-hoc tests and *t*-tests against chance level with *pinguoin*^45^ *ttest* (version: 0.5.5). Reported values represent mean ± standard deviation unless stated otherwise. Statistical results are not corrected; instead, we report multiple alpha thresholds to facilitate comparison to other studies.

### Generative Artificial Intelligence Tools

*Copilot* (Microsoft, Redmond, Washington, USA, version: 2.20260326.20.0) was used selectively to enhance the clarity and precision of the wording in the manuscript.

## Results

### Butterworth filter order does not affect classification performance

Since the Butterworth filter was only used to calculate the envelope biomarkers and the filter order has no effect on their classification results (Figure S 1), we limit further analyses to the 8^th^ filter order. This reduces the multiverse to 2376 paths.

### Classification results are not replicable across datasets

The overall classification performance varied significantly between datasets (*t*_10_=4.760; *p*<.001). Classifiers operating on D1 achieve a mean accuracy of 59.4±9.4% across all paths with a wide performance range from 35.4% to 87.5% mean accuracy for the individual paths. The overall performance of classifiers operating on D2 is at chance level with 49.3±8.8% with mean accuracies for the individual paths ranging from 18.9% to 87.2%. Note that some statistical effects for D2 might not be meaningful when the accuracies perform on average around chance level.

### Artifact removal impacts classification performance

During manual revision of the EEG signals, we found technical artifacts (sinusoidal artifacts at ∼5Hz across channels (Figure S 2 & Figure S 3) and exceptionally high spikes (Figure S 4 & Figure S 5) in D1 that were not removed by the automatic artifact removal. Importantly, those technical artifacts were unequally distributed across groups (before subject removal: MDD: 1 (3.4%) / HC: 12 (42.8%); *p*<.001; after subject removal: MDD: 1 (4.0%) / HC: 8 (34.8%)); *p*=.009). For D1, the artifact removal step has a significant impact only on the overall performance of the classifiers operating on the U-BMs (*F_2,10_*=4.201, *p*=.047), However, biomarker performance of both groups is differentially affected by it (interaction artifact removal x biomarker, Figure 1A; U-BM: *F_10,50_*=10.969, *p*<.001; A-BM: *F_8,40_*=2.423, *p*=.031). Analysis of D2 neither reveals an impact of the artifact removal step per se (U-BM and A-BM: *F_2,10_*<3.006, *p*>.095), yet biomarkers are differentially modulated by the chosen method to remove artifacts (interaction artifact removal x biomarker, Figure 1B: U-BMs: *F_10,50_*=3.295, *p*=.002, A-BMs: *F_8,40_*=2.892, *p*=.012). Because of the bias due to the technical artifacts in D1, the remaining analysis includes only the results of the classifiers operating on the manually revised data, leaving 792 paths.

**Figure 1:**
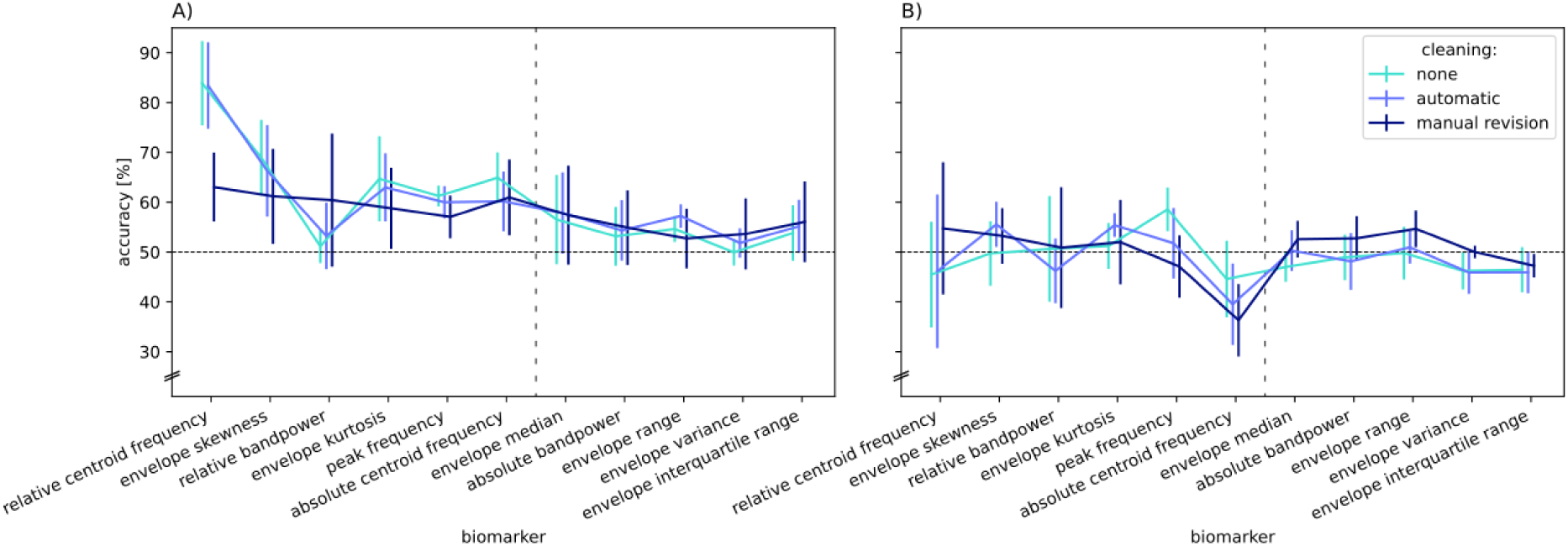
Interaction effects of artifact removal x biomarker on classification performance (balanced accuracy) for A) D1 and B) D2 for 8^th^ Butterworth filter order. Values are averaged across factors for each fold. Error bars depict standard deviations across folds. Biomarkers are grouped by U-BMs (left of the vertical dotted line) and A-BMs (right) and then sorted according to their mean performance across datasets. Abbreviations: D1/2 = dataset 1/2, U-BMs = biomarkers unaffected by normalization, A-BMs = biomarkers affected by normalization.

### Impact of processing steps on classification performance

First, we assess whether the *robustness* of the classification results against the variations of the individual processing steps is *replicable* across datasets. The A-BMs of both datasets are significantly affected by normalization (Figure 2A, D1 (black squares): *F_2,10_*=6.529, *p*=.015); D2 (grey squares): *F_2,10_*=4.534, *p*=.040). Additionally, the performance of the individual biomarkers is differentially modulated by normalization (Figure 3A, D1: *F_8,40_*=3.779, *p*=.002; Figure 3B, D2: *F_8,40_*=3.210, *p*=.006). These main and interaction effects are the only one that both datasets have. However, the pattern of influence differs across datasets. The only other commonality across datasets is that the processing step aggregation affects classification accuracy for neither dataset nor biomarker group (Figure 2C).

**Figure 2:**
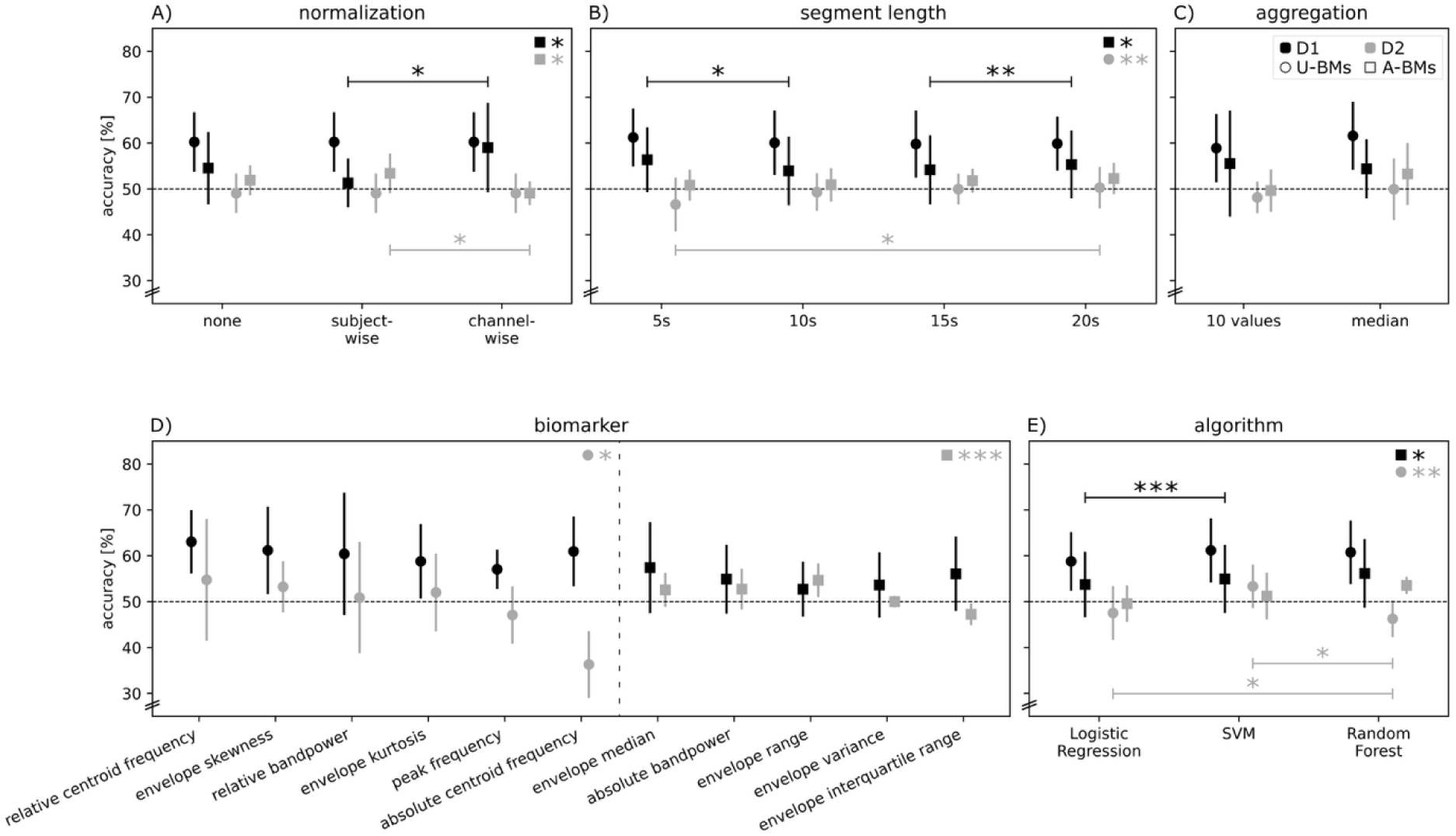
Effects of preprocessing steps, biomarkers, and classification algorithms on classification performance (balanced accuracy) for D1 (black) and D2 (grey) for Butterworth filter order of 8 and manually revised data. Values are averaged across factors for each fold. Error bars depict standard deviations across folds. Main effects for each dataset and biomarker group separate are depicted in the top right corner. Horizontal bars depict post hoc tests. Stars depict an effect of the analyses with *<.05, **<.01, ***<.001, all uncorrected. Biomarkers are grouped by U-BMs (circle markers) and A-BMs (square markers) and then sorted according to their mean performance across datasets. This order is retained for all subsequent figures. Color coding applies to all elements in the figure. Abbreviations: D1/2 = dataset 1/2, SVM = Support Vector Machine, U-BMs = biomarkers unaffected by normalization, A-BM: biomarkers affected by normalization.

**Figure 3:**
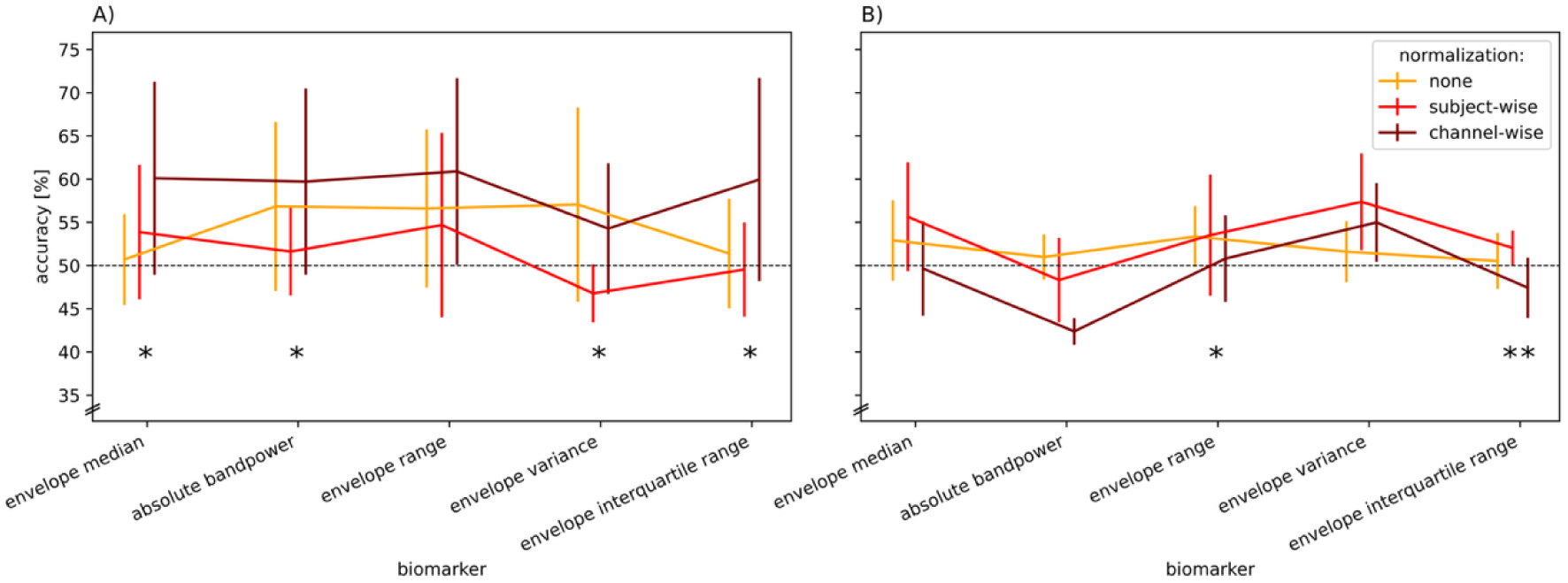
Interaction effects of normalization x biomarker on classification performance (balanced accuracy) for A) D1 and B) D2 for 8^th^ Butterworth filter order and manually revised data only for A-BMs. Values are averaged across factors for each fold. Error bars depict standard deviations across folds. Stars depict an effect of the post-hoc ANOVA for the corresponding biomarker with *<.05, **<.01, ***<.001, all uncorrected. Abbreviations: D1/2 = dataset 1/2, A-BMs = biomarkers affected by normalization

For D1, the performance of classifiers with U-BMs were not affected by any processing steps (all factors *p*>.06). The classification performance of the A-BMs is significantly affected by segment length (Figure 2B, black square: *F_3,15_*=3.522, *p*=.041), and algorithm (Figure 2E, black square: *F_2,10_*=4.606, *p*=.038).

D2 is more severely affected by processing steps. The performance of both biomarker groups is significantly affected by biomarker (Figure 2D, U-BMs (grey circle): *F_5,25_*=3.371, *p*=.018; A-BMs (grey squares): *F_4,20_*=13.102, *p*<.001). Furthermore, the performance of the individual biomarkers is differentially modulated by algorithm (U-BMs: *F_10,50_*=2.777, *p*=.008; A-BMs: *F_8,40_*=4.836, *p*<.001). The performance of the U-BMs significantly depends on segment length (Figure 2B, grey circle: *F_3,15_*=5.567, *p*=.009), and algorithm (Figure 2E, grey circle: *F_2,10_*=9.947, *p*=.004). The performance of the individual A-BMs is differentially modulated by segment length (*F_12,60_*=9.427, *p*<.001). The interaction effects between normalization x algorithm and segment length x aggregation are also unique for the A-BMs of D2 (both interactions *p*<.003).

### Biomarkers are differentially modulated by preprocessing steps

To examine more closely how individual processing steps affect each biomarker, we conducted separate ANOVAs for all biomarkers. Three A-BMs are in both datasets influenced by the same set of processing steps. However, each of them is additionally affected differently by one processing step that differs between datasets. The **absolute band power** is affected by segment length (D1: *F_3,15_*=3.355, *p*=0.047; D2: *F_3,15_*=4.130, *p*=.026) and additionally in D1 affected by normalization (D1: *F_2,10_*=6.236, *p*=0.017). **Envelope range** is affected by segment length (D1: *F_3,15_*=6.057, *p*=0.007; D2: *F_3,15_*=12.772, *p* <.001) and algorithm (D1: *F_2,10_*=4.178, *p*=0.048; D2: *F_2,10_*=5.714, *p*=.022), and additionally in D2 affected by normalization (D2: *F_2,10_*=4.172, *p*=.048). The last one of this group, **envelope interquartile range**, is affected by normalization (D1: *F_2,10_*=5.029, *p*=0.031; D2: *F_2,10_*=14.171, *p*=.001) and additionally in D2 affected by algorithm (D2: *F_2,10_*=7.833, *p*=.009). Furthermore, the three U-BMs **relative centroid frequency**, **relative band power**, and **absolute centroid frequency** are not affected by any processing steps in both datasets (all factors, D1: *p*>.08; D2: *p*>.11).

The remaining five biomarkers were differently affected in D1 and D2. **Envelope median** (A-BM) is affected by normalization in D1 (*F_2,10_*=6.610, *p*=.015), and by segment length in D2 (*F_3,15_*=4.782, *p*=.016). **Envelope variance** (A-BM) is also affected by normalization in D1 (*F_2,10_*=6.567, *p*=0.015) but not affected at all in D2 (all factors, *p*>.09). This pattern is reversed in the remaining three U-BMs, which are not affected in D1 (all factors, *p*>.05). Yet in D2, **envelope skewness** and **envelope kurtosis** are affected by algorithm (*F_2,10_*=14.262, *p*=.001; *F_2,10_*=6.744, *p*=.014; respectively), and **peak frequency** is affected by segment length (*F_3,15_*=4.016, *p*=.028).

### Most biomarkers are not robust against chance level

The smaller the impact of the processing steps on the diagnostic accuracy of a biomarker, the more *robust* it is to analytical variability (smaller range in Figure 4A). The most *robust* biomarkers vary between datasets with **envelope kurtosis** (14.6%) for D1 and **relative centroid frequency** (13.9%) for D2. The least *robust* biomarker is the **envelope range** with 27.6% in D1 and 43.9% in D2.

**Figure 4:**
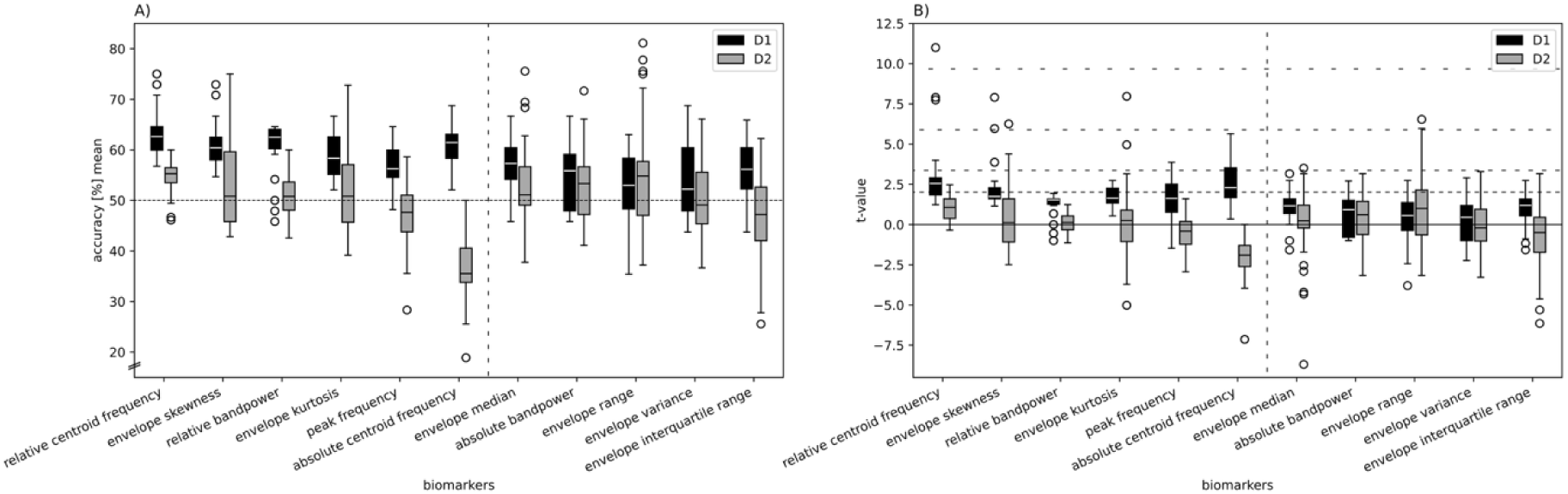
Robustness to analytical variability. A) Classification accuracies (balanced accuracy) across paths (n=72) with Butterworth filter order of 8 and manually revised data. Mean accuracies for each path are averaged across folds. Horizontal dotted line depicts chance level. Vertical dotted line separates U-BMs (left) from A-BMs (right). B) t-statistics for each path of each biomarker (n=72) with Butterworth filter order of 8 and manually revised data. Horizontal dashed lines depict varying degrees of alpha levels from 0.05 (lowest and narrowest dashed line), 0.01, 0.001, to 0.0001 (highest and widest dashed line). Data points are depicted as outliers when their values extend beyond 1.5 times the interquartile range from the first and third quartile, respectively. Abbreviations: D1/2 = dataset 1/2, U-BMs = biomarkers unaffected by normalization, A-BM: biomarkers affected by normalization.

The *t*-statistic combines the performance of a classifier with its *internal replicability*, acknowledging the stochastic nature in the data sampling process (Figure 4B). While inference testing is standard in medicine and neuroscience, biomarker studies from the ML field tend to neglect *replicability* in favor of performance.^46^ Examining the *robustness* of biomarkers within paths from the perspective of a quasi-meta-analysis, we find that only 19.8% of all diagnostic classifiers, i.e., 19.8% of all paths across biomarkers and datasets, perform significantly better than chance level on our most lenient alpha level of 0.05 (Figure 4B lowest dashed line). Along this line, there is no biomarker that surpasses this threshold on all paths. Furthermore, only one biomarker, the **relative centroid frequency** in D1, contains one path (segment length: 15s, aggregation: median, algorithm: SVM) that performs better than our most conservative alpha level (Figure 4B, highest dashed line: *p*<.0001). Notably, just four biomarkers in D1 (**relative centroid frequency**, **envelope skewness**, **envelope kurtosis**, **absolute centroid frequency**) obtain only positive *t*-values, meaning that all their paths perform above chance level. Another interesting behavior show biomarkers like the **envelope kurtosis** in D2, where 83.3% of all paths perform below our most lenient alpha level of 0.05 yet one path (segment length: 20s, aggregation: individual values, algorithm: Random Forest) exceeds our second most conservative level of 0.001.

In order to demonstrate the *replicability* of the performance results within the datasets, we calculated the standard deviations of classification accuracies across folds obtained by the six-fold cross-validation for each path (Figure 5). The lowest median of the standard deviations is 10.8% for **peak frequency** in D1 and 10.6% for **envelope kurtosis** in D2. The highest variability we find for the **relative band power** in D1 (median of std=19.4%) and **absolute centroid frequency** in D2 (median of std=16.8%). However, these variations need to be considered with caution in this study since each test set for cross-validation only contained five to eight subjects, i.e., the resolution for the accuracy ranges between 12.5% and 20%.

**Figure 5:**
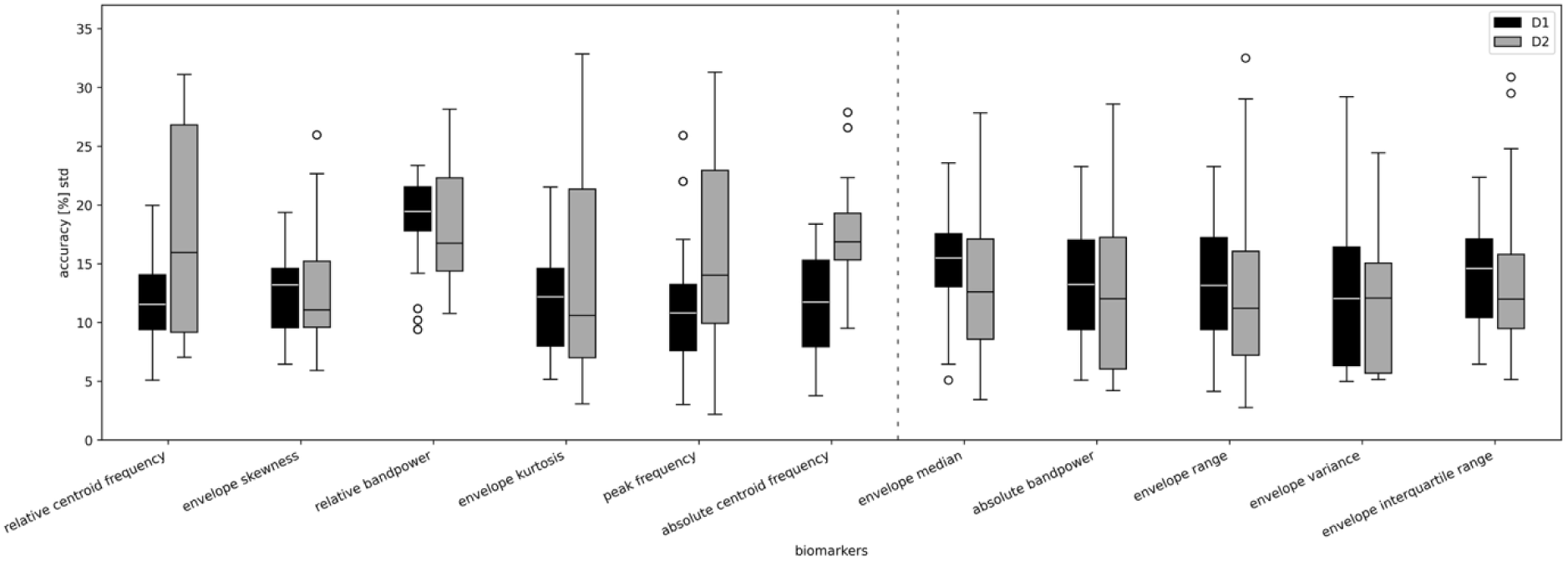
Replicability within datasets. Variability of classification accuracies (balanced accuracy) across folds within each path (n=72) with Butterworth filter order of 8 and manually revised data. Standard deviations across the six folds are shown for each path. Vertical dotted line separates U-BMs (left) from A-BMs (right). Data points are depicted as outliers when their values extend beyond 1.5 times the interquartile range from the first and third quartile, respectively. Abbreviations: D1/2 = dataset 1/2, U-BMs = biomarkers unaffected by normalization, A-BM: biomarkers affected by normalization.

To summarize, we observed four different kinds of performance patterns in the analyses. Mostly *robust* to analytical variability: The biomarker performs significantly above chance-level for most paths (Figure 4B, e.g., D1 **relative centroid frequency**). Lack of *replicability* across datasets: The biomarker performs above chance level in one dataset and below in the other (Figure 4A, e.g., **absolute centroid frequency** (*t_71_*=31.973, *p*<.001). Limited *replicability* within datasets: The performance within a dataset and path shows high variance across folds (Figure 5, e.g., D2 **envelope kurtosis**). Not *robust* to analytical variability: The biomarker performance diverges widely within a dataset depending on the chosen processing path (Figure 4A, e.g., D2 **envelope range**).

### Biomarker patterns

Another perspective on the impact of the processing steps on the biomarkers is presented by a correlation analysis between the different paths. The focus here is on the biomarker patterns – that is, whether the biomarkers exhibit similar or divergent performance relative to one another.

The correlations are in general higher within (Figure 6 and Figure S 8) than across datasets (Figure S 9). Channel-wise normalization of D2 seems to align the classification patterns to the ones of D1. Furthermore, 5s segments and channel-wise normalization (in D1 as well as in D2) increase correlation between datasets.

**Figure 6:**
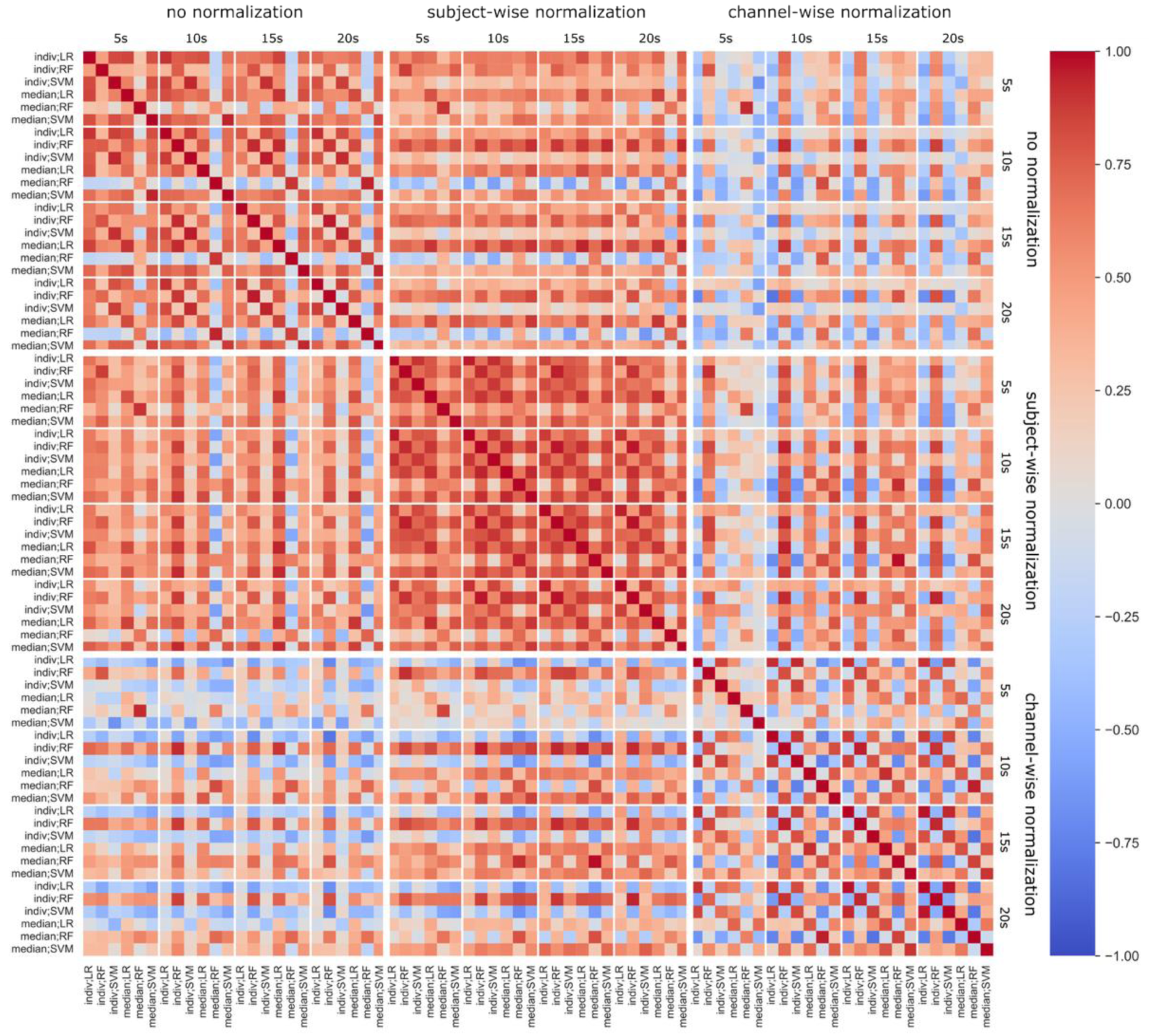
Correlation of classification performance (balanced accuracy) of paths in D1 with Butterworth filter order of 8 and manually revised data (n=72). Path names are constructed as follows: aggregation; algorithm. Big squares depict normalization variants; small squares depict various segment lengths. Abbreviations: LR = Logistic Regression, RF = Random Forest, SVM = Support Vector Machine.

For D1, normalization variants exert distinct effects on the resulting biomarker patterns (Figure 6). Paths employing subject-wise normalization exhibit high correlations without marked differences across the other factors. Paths without normalization also show a high degree of internal consistency, except for those combining median as aggregation step with Random Forest as algorithm. In these cases, the biomarker patterns diverge from the other non-normalized paths. Under channel-wise normalization, we observe that paths using Random Forest display biomarker patterns that are less consistent with the remaining pipelines than those using alternative algorithms.

The raw values of the classification results can be found in the supplementary material (“metrics.csv”) for analytical reproducibility.

## Discussion

This study assesses two types of reproducibility^7^ across processing pipelines for alpha band EEG biomarkers used in a diagnostic task for MDD detection. It evaluates the *robustness* to analytical variability of those biomarkers by assessing their diagnostic performance across a multiverse of possible processing pipelines, as well as their *replicability* with two separate publicly available datasets. Including only linear biomarkers from the EEG alpha band, we achieve diagnostic accuracies that range from 81% down to chance level depending on the dataset, preprocessing variations, biomarkers, and classification algorithm applied. Even though the datasets used in this study are rather small, they are the most widely analyzed publicly available datasets for EEG biomarker research in MDD.^28^ More than a third of studies in this field in the last five years base their findings on these datasets, more than 75% of the studies based on public datasets use these data and from those studies, more than 75% use them as standalone datasets without further data to *replicate* their findings.

### Replication of findings with two publicly available datasets

*Replicability* of biomarker research is important for generalizability and application in a clinical setting. The *replication* of findings across datasets was largely not possible in this study despite their harmonization. It was severely hampered by the classifiers trained on D2 performing on average only on chance level. Similar findings have been reported before when working with these datasets; D2 shows either a behavior contrary to other datasets analyzed^26^ or a performance around chance level for approaches that work with other datasets^13^. Limited *replicability* across datasets despite the same analysis pipeline might have sources related to the heterogeneity of the disease, differences in the study design, or in the data acquisition.

A multifaceted disorder such as MDD is heterogeneous in terms of severity and symptoms, progression, (drug) treatment (success), and the presence of comorbidities.^47^ Sample sizes of overall 30 to 50 participants^6^ are very likely too small to capture the variability observed in MDD. Furthermore, the label MDD is assigned on various criteria, including clinical diagnoses based on the DSM-IV manual or by sum-scores of questionnaire-based assessments,^26^ which cannot reflect the prevailing heterogeneity of the condition. This holds for both patients and HCs, labels that are in fact too simple for capturing the complex physiology and phenology of a disorder such as MDD.^48^ As a consequence, the effect sizes of studies are often inflated,^7,49,50^ leading to overestimation of the selectivity of the biomarkers studied. The patients of both datasets in this study have been diagnosed based on DSM-IV criteria but cannot be compared with regard to other clinical factors since information about them is missing for one or both datasets. Given the small sample sizes in this study, patient variability both within and across datasets may not have been adequately captured, likely contributing to the limited *replicability*. In order to understand the physiological underpinnings of a heterogeneous disease like MDD and find reproducible biomarkers, we need to characterize subjects much better. The variability in symptoms and their expression as well as comorbidities need to be assessed thoroughly and represented in the data.^10^ The better characterized the datasets are, the higher the chances that data can also be pooled across studies. Complementary, dedicated studies to systematically assess these impacting factors are needed. Along the same line, the study populations need to be well characterized with additional information, e.g., demographic data or other factors about the current state of the subjects. The datasets for this study were collected in different countries and the patients from D1 were on average about ten years older. These differences might have further contributed to the lack of *reproducibility*.

Comparability of studies may further be hampered by the lack of standardization in the recording of EEG data. There are differences in the recording setup, electrode placement, and electrode referencing. Technical differences like the electrode placements, e.g., the quasi-standard of the EEG-10-20/10/5 system^32^ or the Geodesic Sensor Net configuration^33^ might be other sources of variability. It has been shown that even electrodes from the same landmark-standardized placements record different underlying brain regions between subjects,^51^ questioning the comparability of subjects as well as different electrode placements. More variability in technical recording properties comes from the choice of the electrode type, sampling frequency, and acquisition hardware. Moreover, settings like the time of the day, temperature of the room, or resting-state conditions can introduce further variability. Obtaining robust neurophysiological representations^52,53^ across all these variations requires data sharing, and collaborative efforts are sorely needed.^7^

*Replicability* cannot just be assessed across datasets, but also within. To assess the *replicability* within datasets, we used six-fold cross validation to receive a range of classification results instead of randomly drawing one single training- and test-set. This revealed high fluctuations of classification accuracy even within datasets, confirming the notion of high heterogeneity within the groups MDD and HC. Yet, these findings are limited by the small sample size leading to five to eight subjects per test group, which means 2 to 4 subjects per group (MDD/HC). One falsely classified subject can thus lead to an accuracy drop of 12.5% to 20%. These measures of variability should therefore not be over-interpreted and serve merely as an illustration what kind of statements would be possible with this approach.

### Robustness to analytical variability

The vast variability in processing and analysis pipelines for neuroimaging data is another potential source for lack of reproducibility, which is addressed with meta- and multiverse-analyses, or multi-analyst studies.^8^ Performing a multiverse analysis is advantageous when multiple equally valid processing options are available. If a finding is rather *robust* across the multiverse space this can strengthen the credibility of this finding. If it is not, it might resolve discrepant previous findings. With our multiverse analysis we find a marked influence of some of the investigated processing steps.

The most notable effect on the classification was introduced by data cleaning. Manual revision of D1 significantly reduced the performance of some biomarkers, especially of the group unaffected by normalization (U-BM). Insights from the revision process reveal technical artifacts (spikes and 5 Hz-noise) in this dataset that are not captured by automatic ICA-based artifact detection algorithms like ICLabel and that are unequally distributed across diagnostic groups. It is therefore recommended to always review the data after automatic artifact removal. Even though D1 is frequently used^28^, we did not find a study that commented on this problem. From the studies included in a recent review^28^, nine of them report methods not suitable for removing these specific artifacts, and three of them did not mention those type of artifact despite performing manual artifact removal. Although we removed those artifacts, our approach might introduce a bias since we excluded segments with artifacts in all paths, solely examined through the manual revision. With this approach we, therefore, remove detected artifacts also in paths where these artifacts would not have been removed.

Normalization has a significant influence in both datasets, yet with opposing effects. Interestingly, in D1 channel-wise normalization outperforms subject-wise normalization, even though this destroys the spatial patterns across electrode locations. Given the performance of D2 around chance level, we refrain from interpreting effects here even when they are statistically significant. The impact of segment length has been shown to influence classification performance,^15^ a finding we partially *replicate,* yet not consistently. The tendency for better performance with longer segments suggests that the discriminatory features of MDD may be embedded in the course of time. Nonetheless, dividing recordings into sections of such length reduces the amount of data available for training; therefore, this processing choice is always a trade-off. Random Forest performs better overall in D2, yielding paths for biomarkers to perform robustly with a high accuracy. This indicates that the data space in these cases is not as well linearly separable since the other two algorithms are linear classifiers. While this study provides the multiverse analysis across six processing steps, further variations to them and further steps in general such as feature extraction and selection as well as evaluation of the models offer room for further variations and need to be considered in future research.

### Biomarkers

The performances of both datasets were significantly impacted by the biomarker choice, yet “good performing biomarkers” vary substantially between datasets and processing pipelines. If a “good pipeline” was accidentally chosen in a traditional study only implementing one pipeline, a biomarker performing well in this pipeline would be reported as a good EEG biomarker for MDD even though it might perform badly with all other processing options. Conversely, a biomarker with a bad performance in one pipeline might be disregarded for further analyses despite its potential, even though a specific processing choice might explain the bad performance.

Even though the explanatory power of our analysis is limited by the size of the datasets, we find some biomarkers whose pattern demonstrate different types of robustness that are methodologically interesting. **Absolute centroid frequency** is in D1 very robust to analytical variability with classification performances above chance level across all pipelines, presents a high within-dataset replicability, and some paths with excellent performance. Even replication with D2 is possible to a certain degree. Similarly, **envelope skewness**, **envelope kurtosis**, and **absolute centroid frequency** exhibit a similar pattern for D1 but they fail to replicate completely in D2. In contrast, we observe that some of the biomarkers that perform on chance level on average have an acceptable performance and *robustness* in individual pipelines, e.g., the **envelope range** in D2. Those type of analyses clearly demonstrate the power of the multiverse approach and provide the opportunity to further reproducibility and translatability of biomarker research.

The set of biomarkers chosen for this study constitutes only a small fraction of possible EEG biomarker. Furthermore, the characteristics of the alpha band exhibit substantial inter-personal variations that are amongst other factors associated with age and cognitive function.^54^ A possible mitigation of these inter-individual differences might be to define alpha not by fixed frequency boundaries but rather by taking the individual alpha peak(s) into account.^55^ Additional biomarkers can be derived from other frequency bands, or come from different categories such as nonlinear biomarkers, connectivity features, or entropy measures and provide complementary characterization of aberrant brain activity in MDD.^3^

## Conclusion

To conclude, this study demonstrates the large influence of choice of processing steps and their combinations for two publicly available EEG resting-state datasets. A multiverse approach in analyses of EEG resting-state data is therefore recommended, at least for the processing steps where an informed decision about a specific option cannot be made. The restricted *replicability* of our findings with two datasets mirrors the inconsistencies in the field very well and highlights the necessity for large and well-curated EEG datasets for MDD research. Along this line, this study is based on rather small datasets, which renders our findings primarily a methodological showcase restricted in generalizability. However, given the frequent use of these two datasets and the fact that this size of dataset is well within the range of data used for current MDD biomarker studies^28^ renders the current study a wake-up call for critical evaluation of the state of the field.

## Data Availability

All data produced in the present work are contained in the manuscript or are available upon reasonable request to the authors.

## Data and Code Availability Statement

Code for biomarker calculation is available at: “calculate_biomarkers.docx”

Raw accuracies of diagnostic classification models are available at: “metrics.csv”

## Supplementary Material

**Figure S 1:**
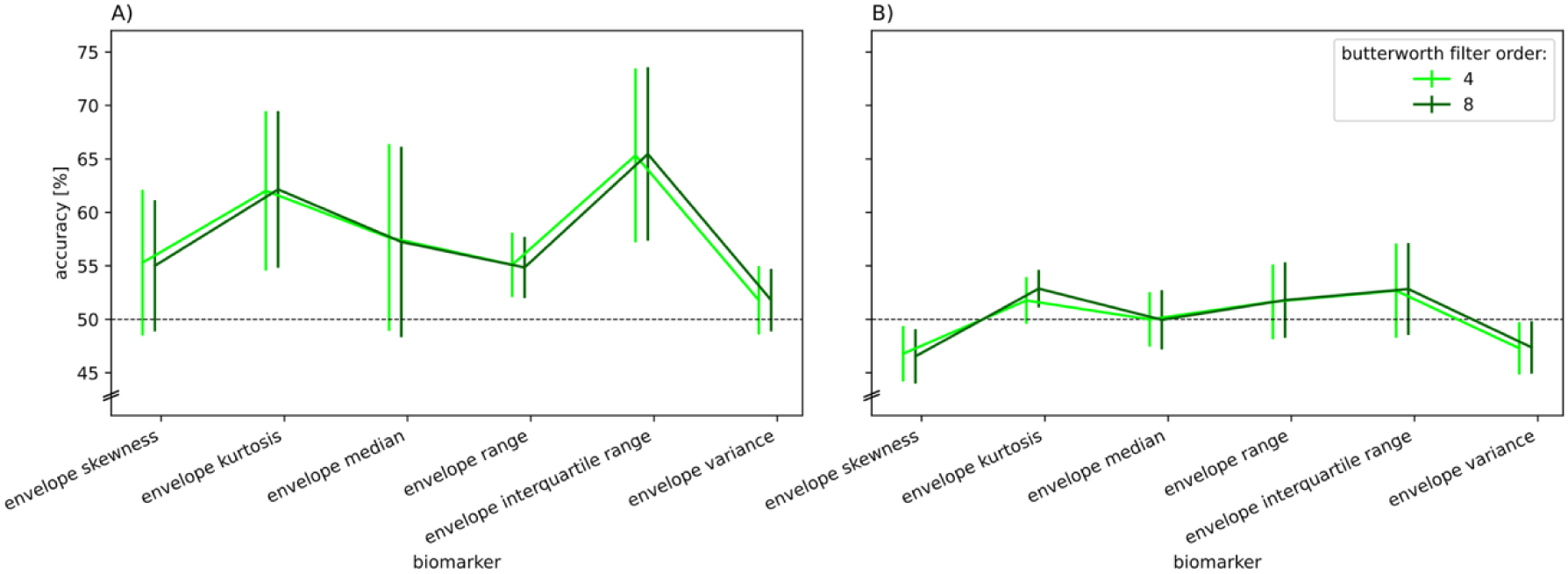
Interaction effects of butterworth filter order x biomarker for A) D1 and B) D2 only for affected envelope biomarkers. Values are averaged across factors for each fold. Error bars depict standard deviations across folds. Abbreviations: D1/2 = dataset 1/2

**Figure S 2:**
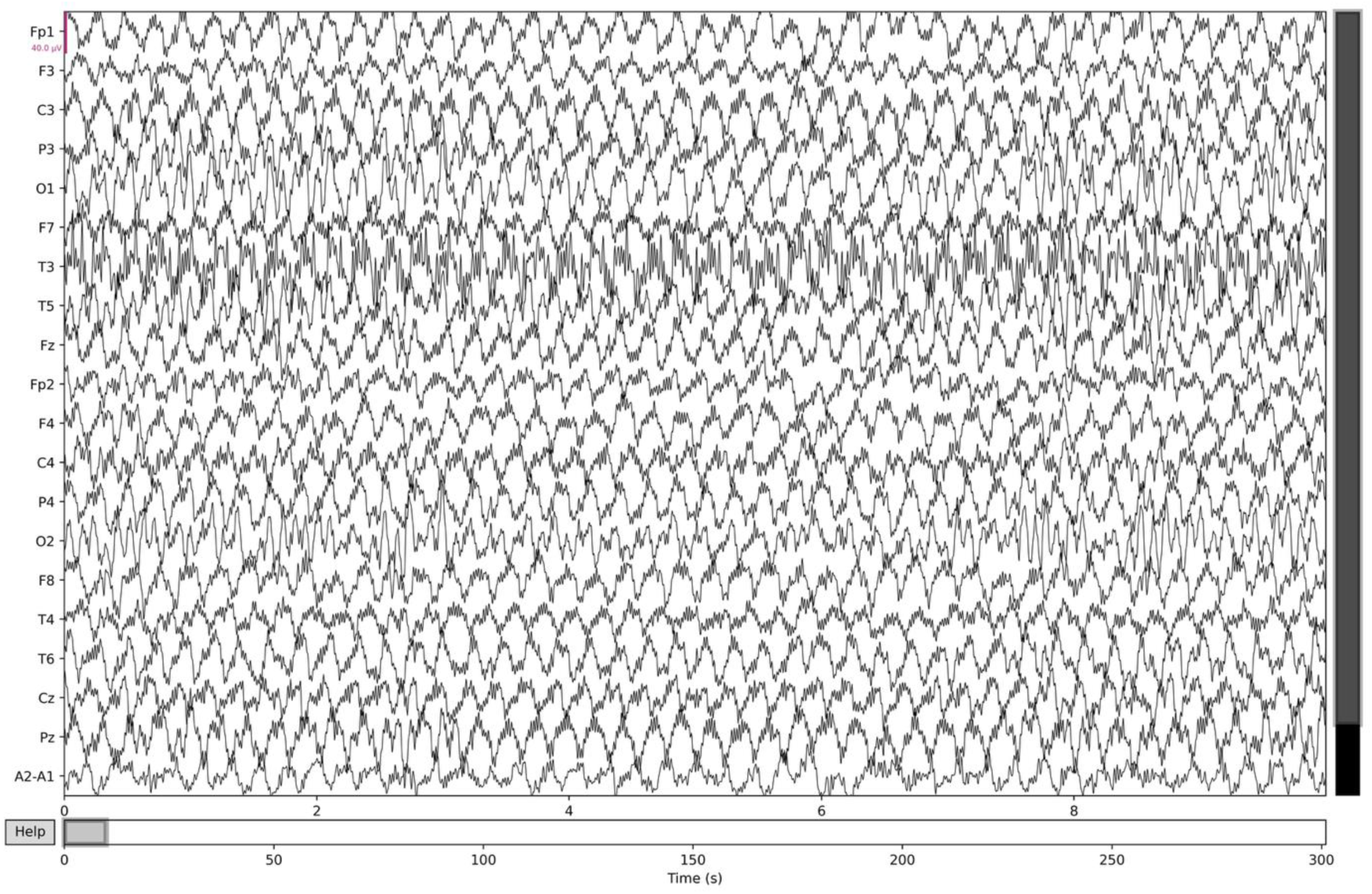
Example of sinusoidal artifacts in D1, 10s window.

**Figure S 3:**
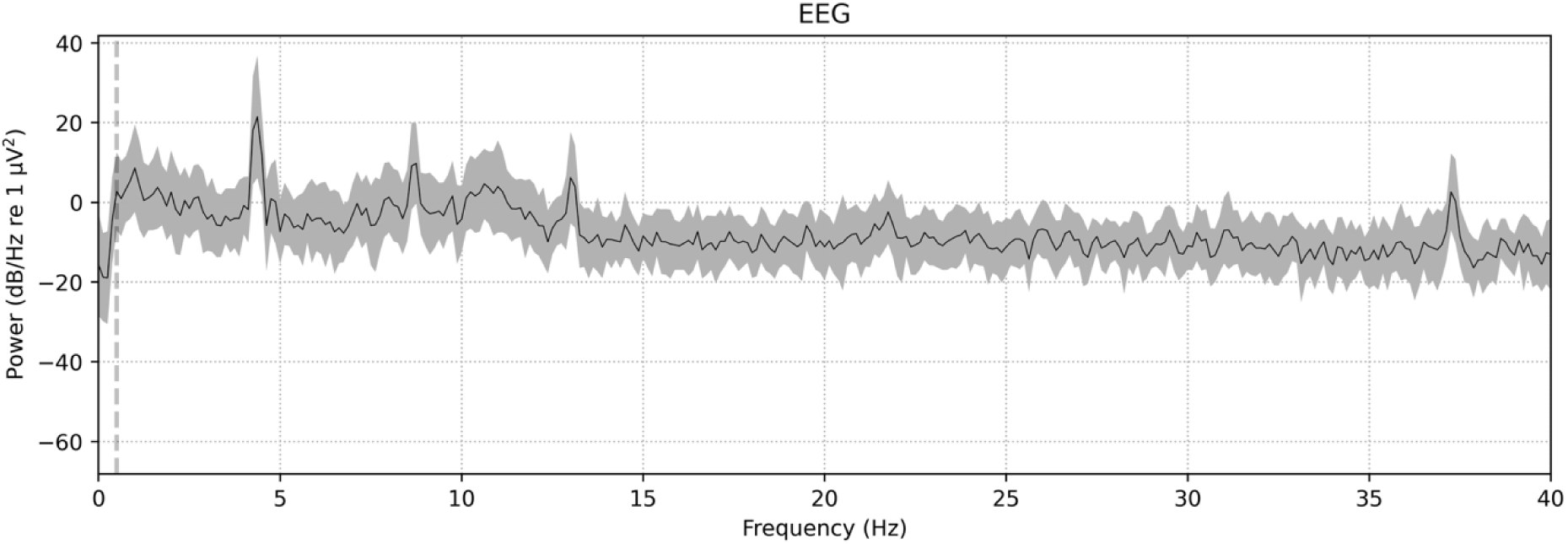
Frequency spectrum corresponding to the window in Figure S 2, averaged across all channels.

**Figure S 4:**
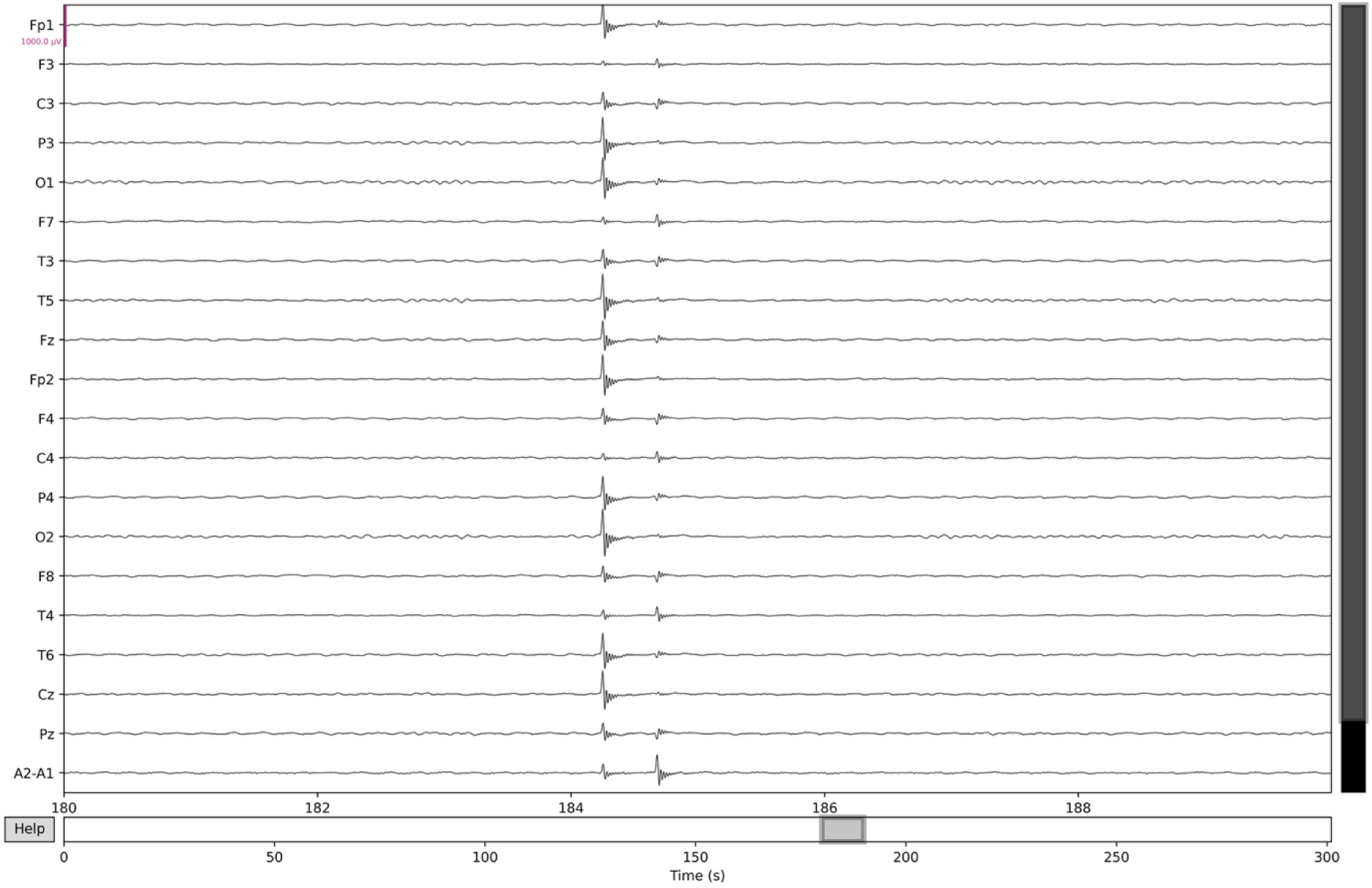
Example of spike artifacts in D1, 10s window. Note that the scaling of the y-axis is 25x larger than the scaling of Figure S 2.

**Figure S 5:**
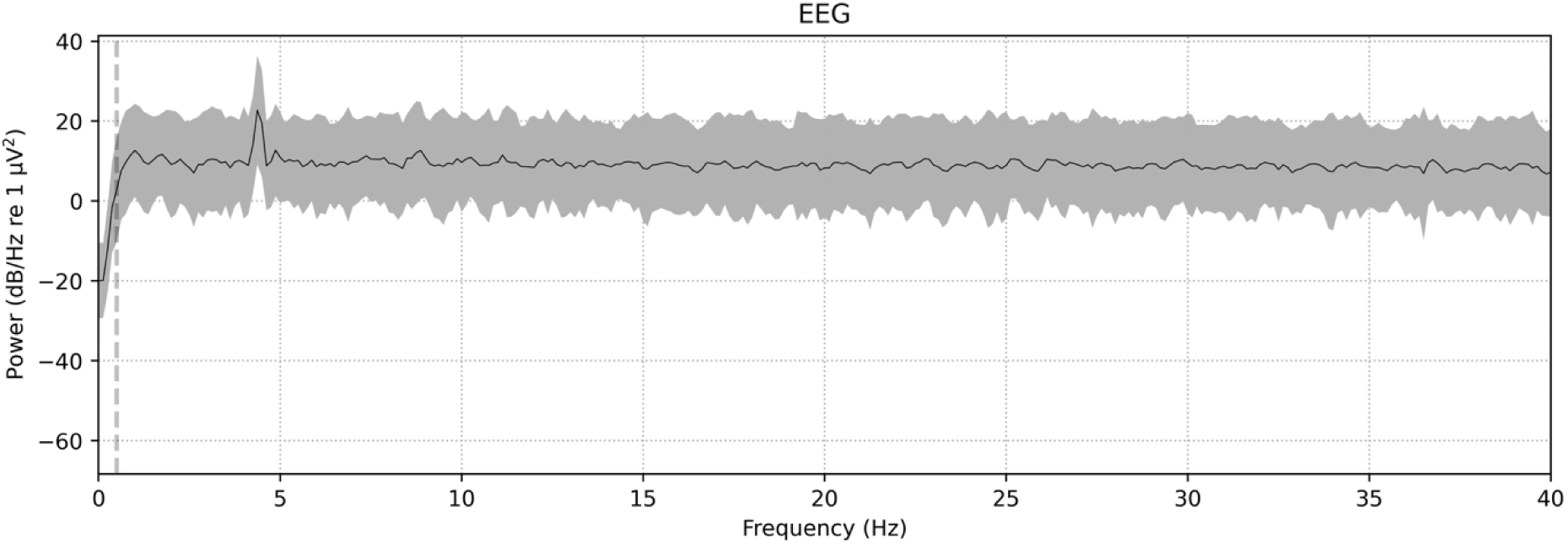
Frequency spectrum corresponding to the window in Figure S 4, averaged across all channels.

**Figure S 6:**
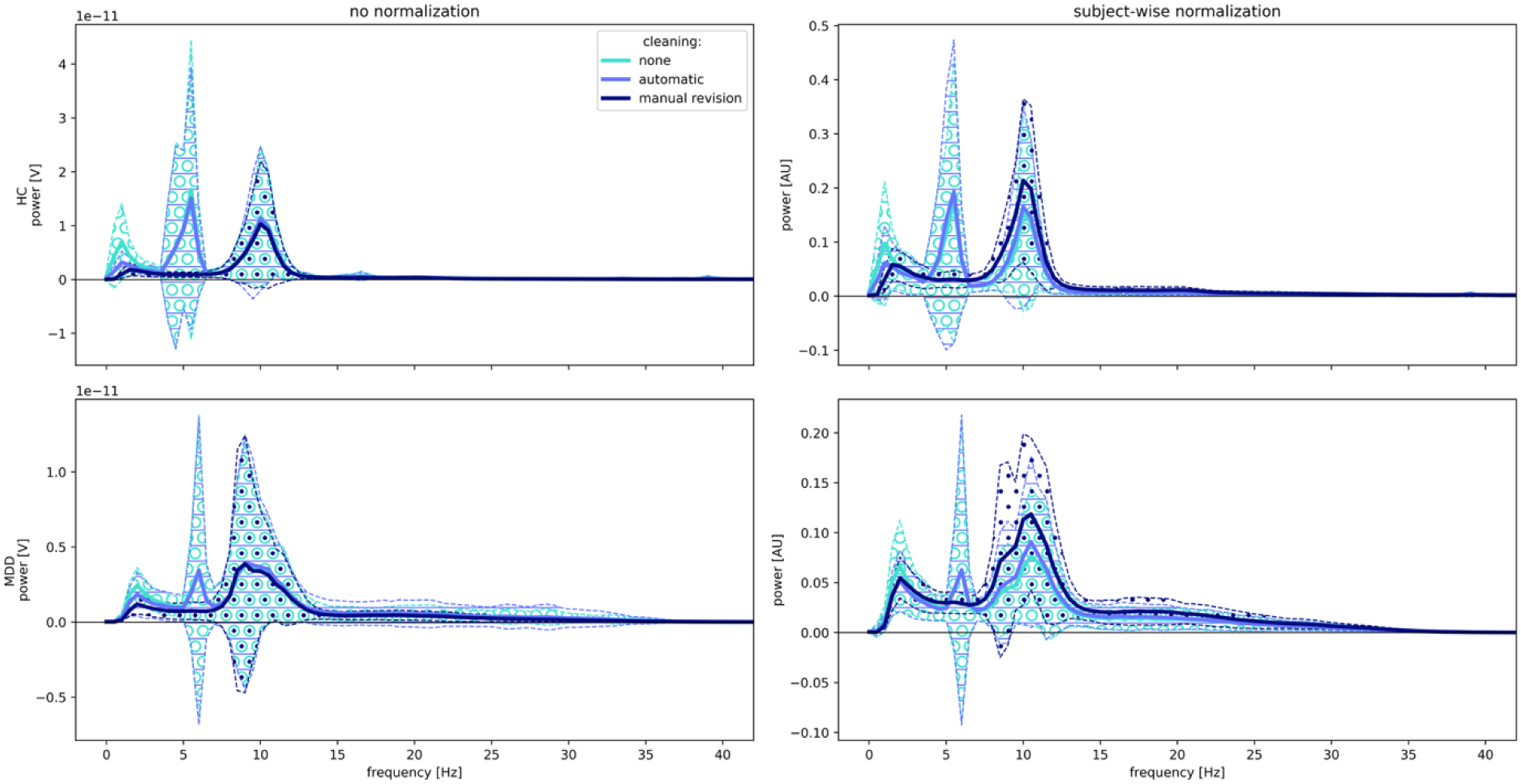
Frequency spectrums of subjects from D1 (HC n=23, MDD n=25) with three artifact removal methods applied (colored lines) for no normalization (left panels) and subject-wise normalization (right panels). Note the different scaling of the y-axis in the four panels.

**Figure S 7:**
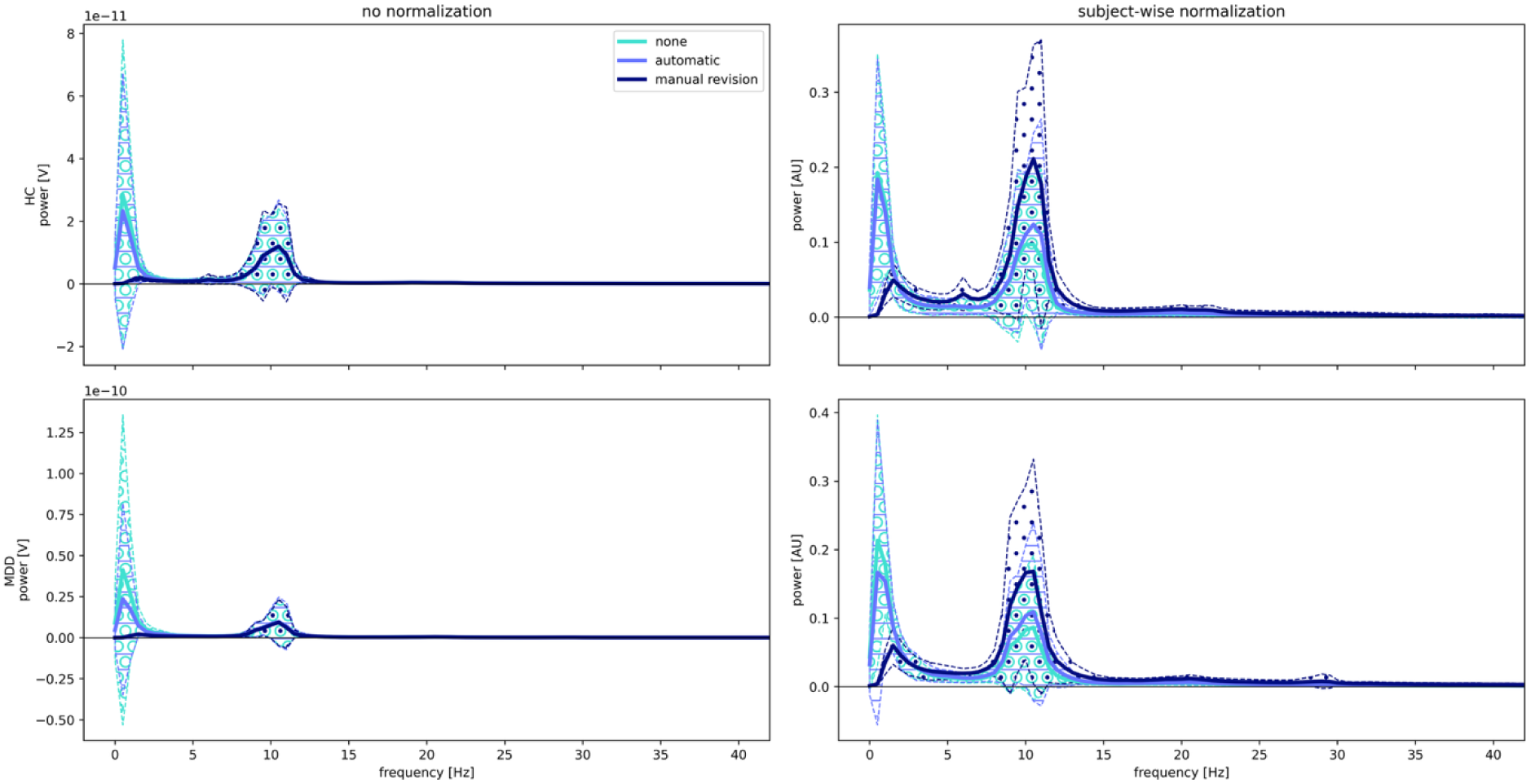
Frequency spectrums of subjects from D2 (HC n=14, MDD n=18) with three artifact removal methods applied (colored lines) for no normalization (left panels) and subject-wise normalization (right panels). Note the different scaling of the y-axis in the four panels.

**Figure S 8:**
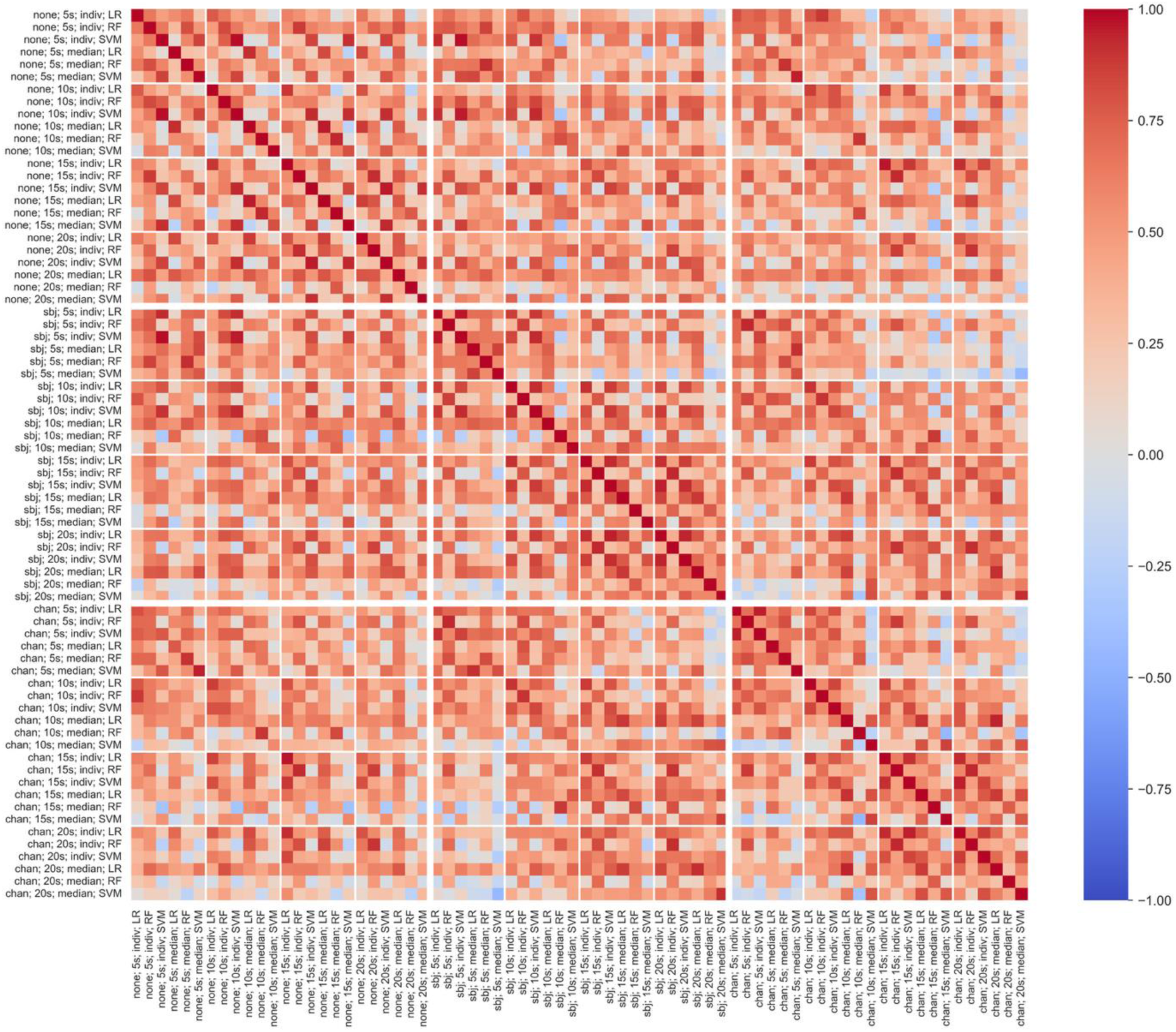
Correlation of classification performance (balanced accuracy) of paths in D2 with Butterworth filter order of 8 and manually revised data (n=72). Path names are constructed as follows: normalization; segment length; aggregation; algorithm. Big squares depict normalization variants; small squares depict various segment lengths. Abbreviations: none = no normalization, sbj = subject-wise normalization, chan = channel-wise normalization, indiv = individual segments, LR = Logistic Regression, RF = Random Forest, SVM = Support Vector Machine.

**Figure S 9:**
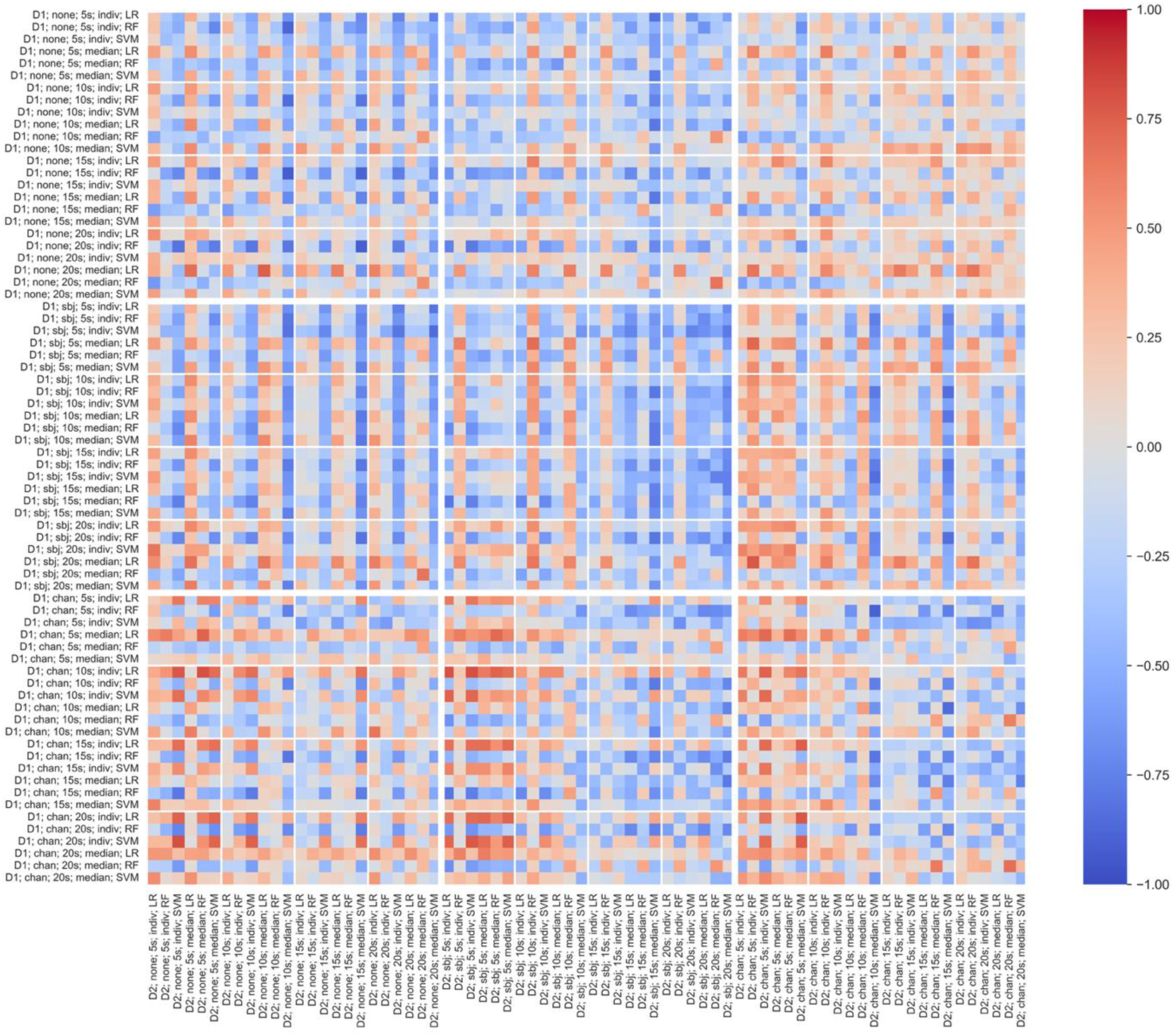
Correlation of classification performance (balanced accuracy) between paths of D1 and D2 with Butterworth filter order of 8 and manually revised data (n=72). Path names are constructed as follows: dataset; normalization; segment length; aggregation; algorithm. Big squares depict normalization variants; small squares depict various segment lengths. Abbreviations: D1/2 = dataset 1/2, none = no normalization, sbj = subject-wise normalization, chan = channel-wise normalization, indiv = individual segments, LR = Logistic Regression, RF = Random Forest, SVM = Support Vector Machine.

